# Does *Toxoplasma gondii* infection have a causal impact on human psychopathology? A Mendelian randomization analysis

**DOI:** 10.1101/2025.11.27.25341156

**Authors:** Jinyong Pang, Matthew J. Valente, Kami Kim, Xiaoming Liu

**Affiliations:** College of Public Health, University of South Florida, Tampa, FL, USA; Morsani College of Medicine, University of South Florida, Tampa, FL, USA

## Abstract

*Toxoplasma gondii* (*T. gondii*) is a prevalent zoonotic parasite that has been implicated in influencing human psychiatric disorders and risk-taking behaviors. Using genome-wide association study (GWAS) data, we conducted two-sample Mendelian randomization (MR) analyses to investigate the potential causal relationship between T. gondii infection and the aforementioned psychiatric disorders and risk-taking behaviors. Contrary to previous epidemiological evidence, our MR analyses do not support a significant causal association between *T. gondii* infection and the studied psychiatric disorders or risk-taking behaviors. These findings challenge the hypothesis that chronic *T. gondii* influences human psychopathology and behavior.

## Introduction

Infection by the protozoan parasite *Toxoplasma gondii* (*T. gondii*) affects approximately one-third of the global human population^1^. This intracellular parasite relies on members of the cat family as definitive hosts for sexual reproduction. All warm-blooded animals can potentially serve as intermediate hosts. Uncooked or undercooked food, as well as water and soil, poses high risks for *T.* gondii infection in humans. These sources may be contaminated by oocysts originating from the feces of infected felines. Once *T. gondii* invades the bodies of intermediate hosts through ingestion, it can either maintain a state of duplication or enter a latent stage. These latent stages are thought to persist indefinitely and may contribute to the alterations of host hormones, neurotransmission within the brain, and neuro-inflammation^2^ providing a mechanism by which persistent *T. gondii* infection or inflammation induced by latent infection could impact neuropsychiatric disease and the host’s behavior. *T. gondii* infection of rodent intermediate hosts influences the behavior of the infected mice, increasing the likelihood of being captured by cats and thus favoring reproduction and transmission^3^.

There is also supporting evidence indicating that latent toxoplasmosis may mentally and psychologically induce behavioral changes in humans^4^. Over the past twenty years, multiple studies focusing on the connection between *T. gondii* infection and various psychiatric disorders, addiction, bipolar disorder, obsessive-compulsive disorder, and Schizophrenia have shown significant associations of OR with the seropositivity of anti-Toxoplasma antibodies, excluding major depression^5,6^. Moreover, links between infection and *T. gondii* and risk-taking behaviors, predominantly encompassing suicidal behavior and risky driving behavior, have been reported^6,7^.

Although accumulating epidemiological evidence has demonstrated an association between *T. gondii* infection and psychiatric disorders or risk-taking behavior using ORs or adjusted ORs, it still remains unknown whether *T. gondii* infection has a causal effect on psychiatric disorders or risk-taking behavior. To answer this question, in this paper, two-sample Mendelian randomization (MR) analyses were performed to estimate the causal association between presumed exposure-outcome pathways and genome-wide association study (GWAS) data utilizing single nucleotide polymorphisms (SNPs) as instrumental variables (IVs)^8^. In this study, four psychiatric disorders, addiction, bipolar disorder, obsessive-compulsive disorder, Schizophrenia, and risk-taking behavior, were adopted as outcomes in MR analysis.

## Results

A total of 25 and 76 SNPs were selected as IVs for anti-*T. gondii* IgG Seropositivity and anti-*T. gondii* IgG levels, respectively (S1-S2 Table). For both the anti-*T. gondii* IgG measures, across all psychiatric disorders and risk-taking behavior, no statistically significant positive causal effect was found (Table 1, S2-S37 Figure). The lack of statistical significance was not likely due to the lack of statistical power, as among the total 36 two-sample MR tests, only two (one dataset for Anti-*T. gondii* IgG Seropositivity on addiction and one for Anti-*T. gondii* IgG Seropositivity on bipolar disorder) have a statistical power < 80% (Table 1). For all the tests, no significant heterogeneity or pleiotropy was detected (S2-S37 Figure, S3-S8 Table). Leave-one-out analyses with sensitivity tests revealed that the effect estimation was not influenced by any dominant SNP (S2-S37 Figure D).

**Table 1.**
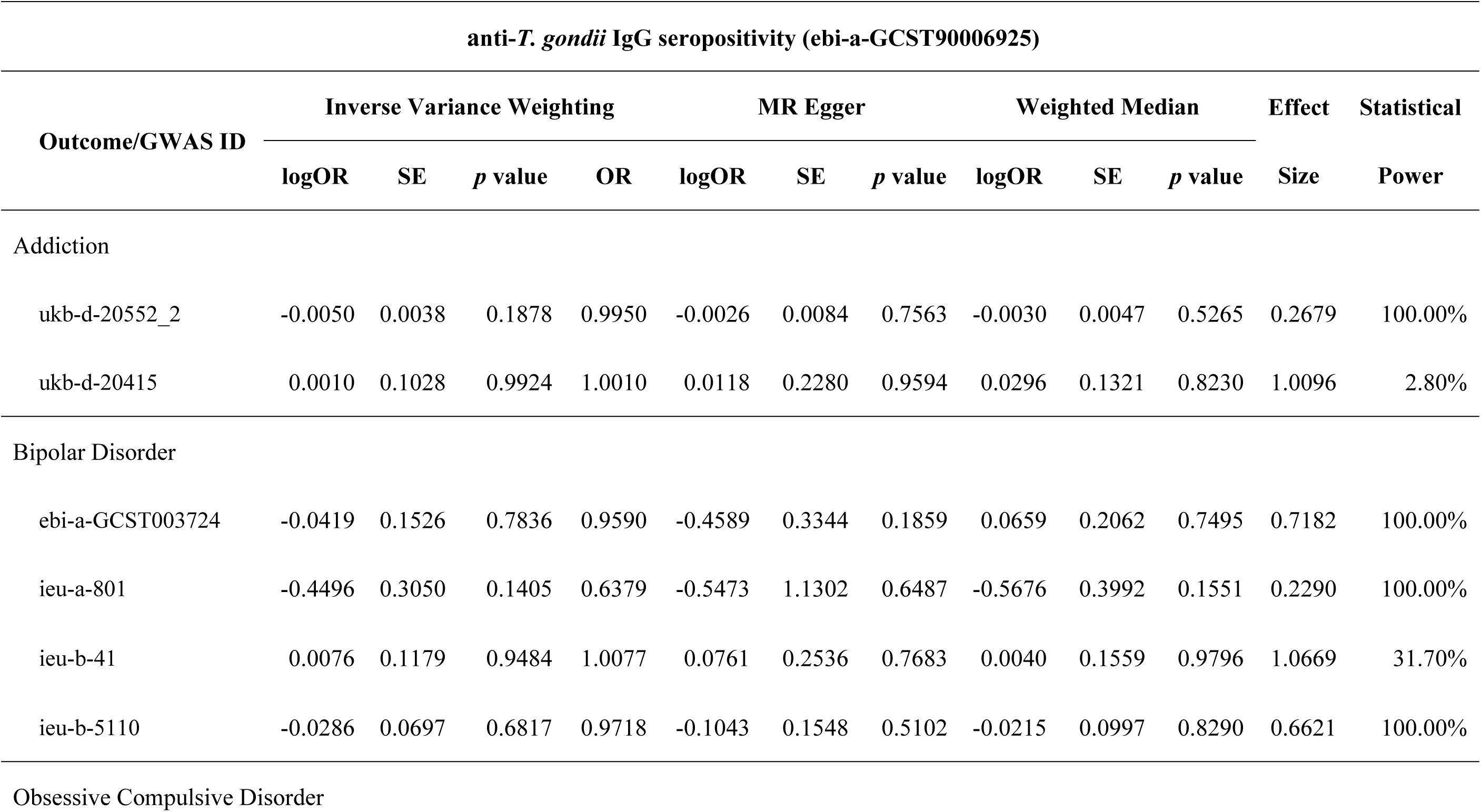
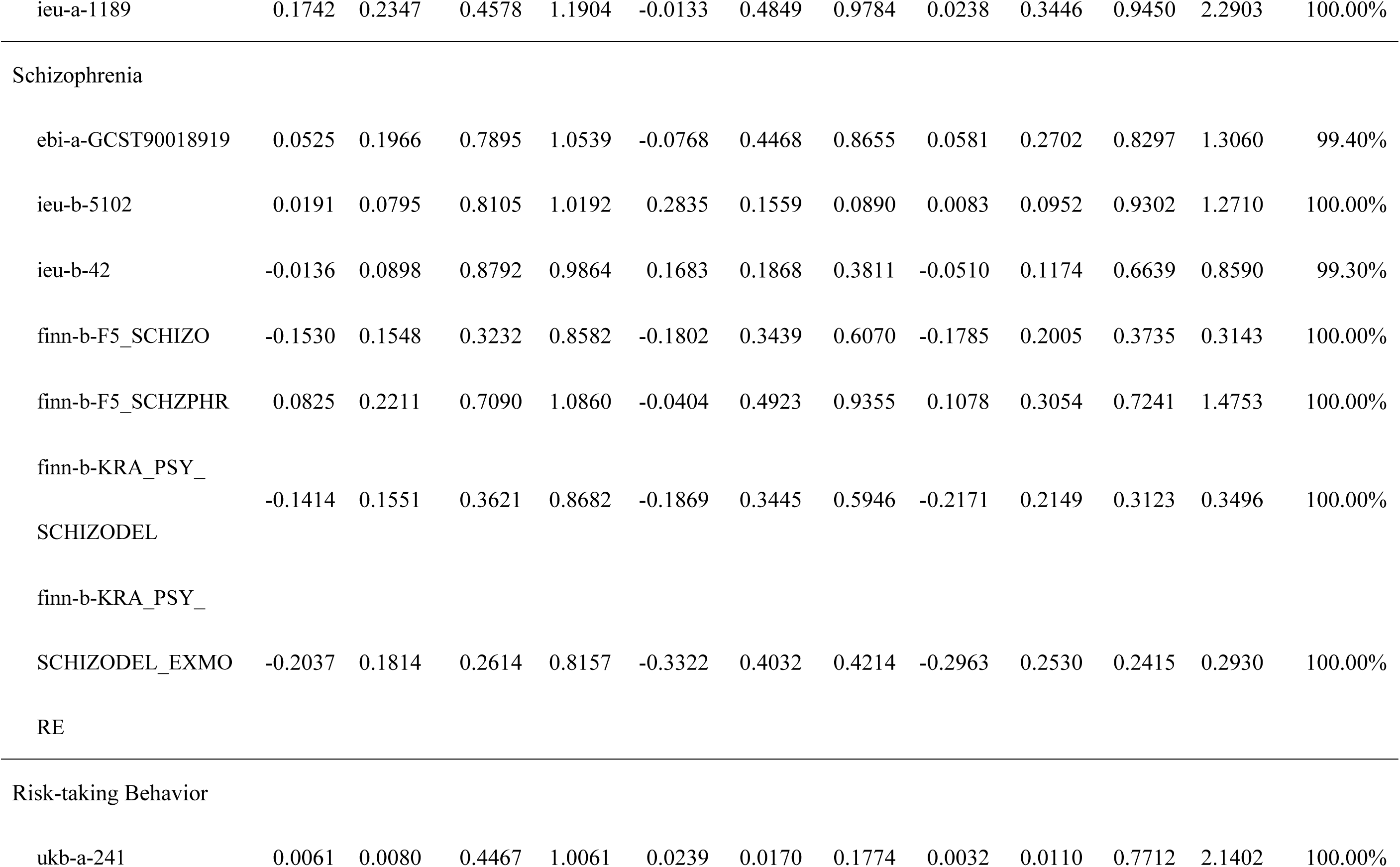
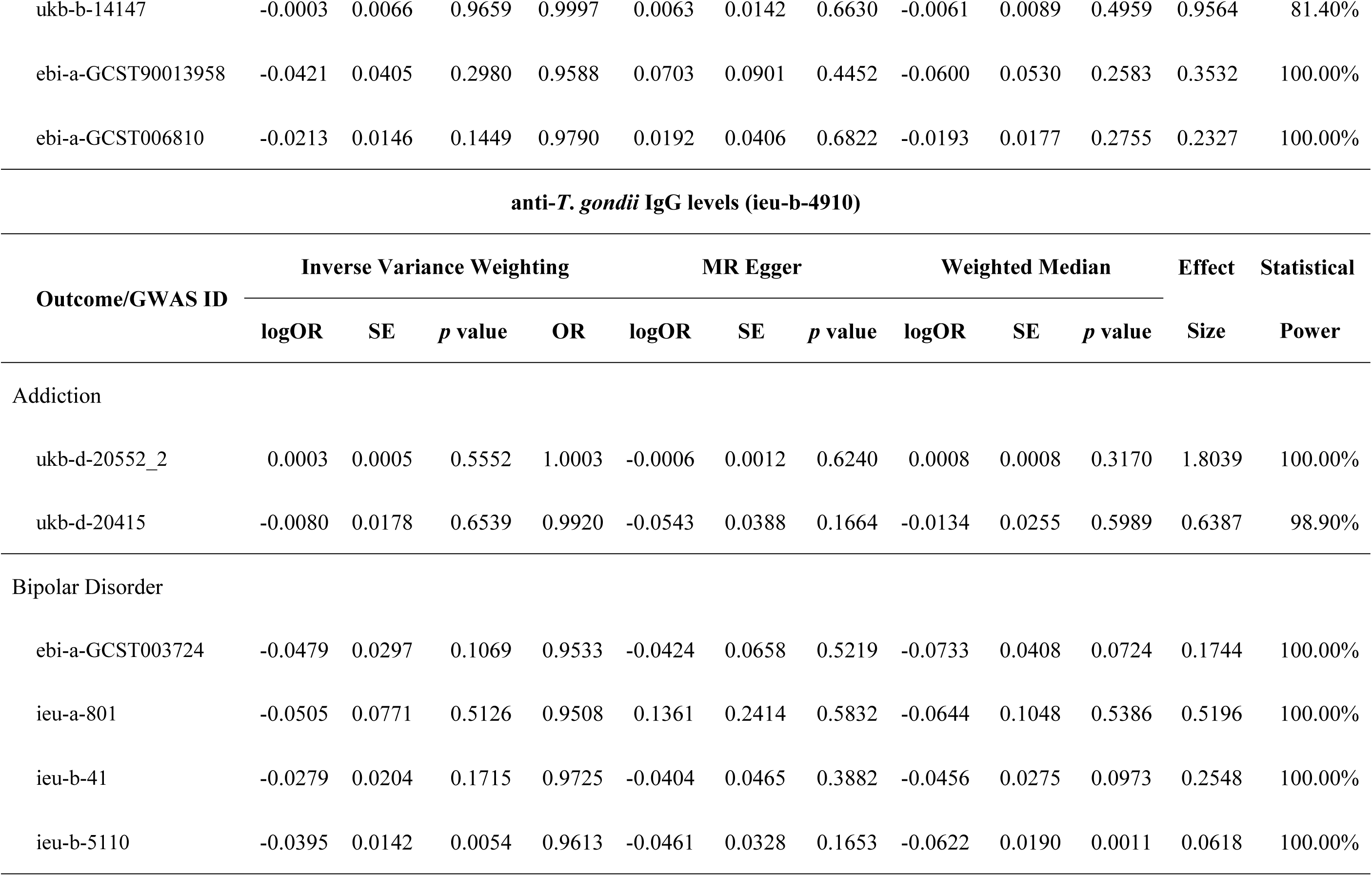
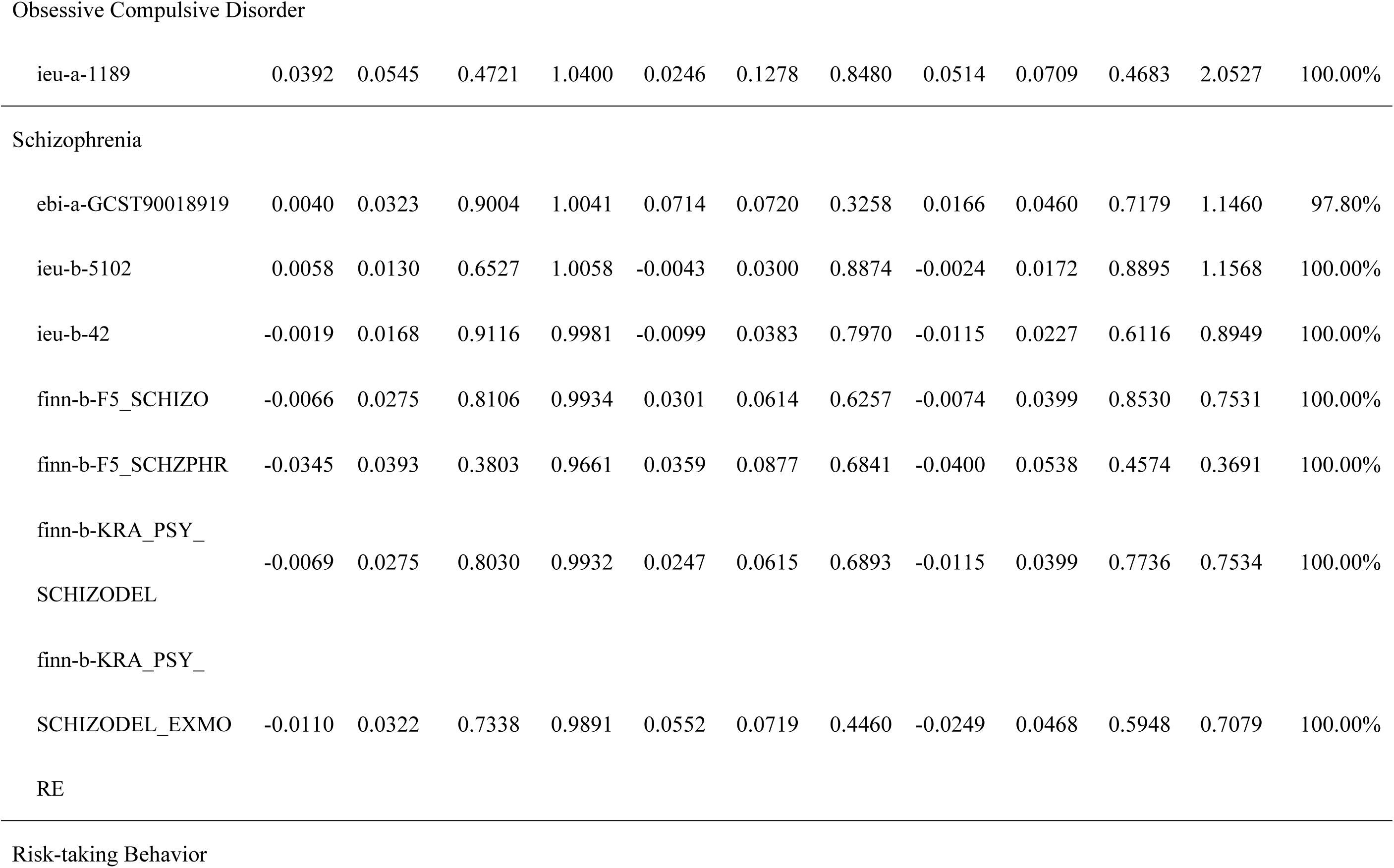
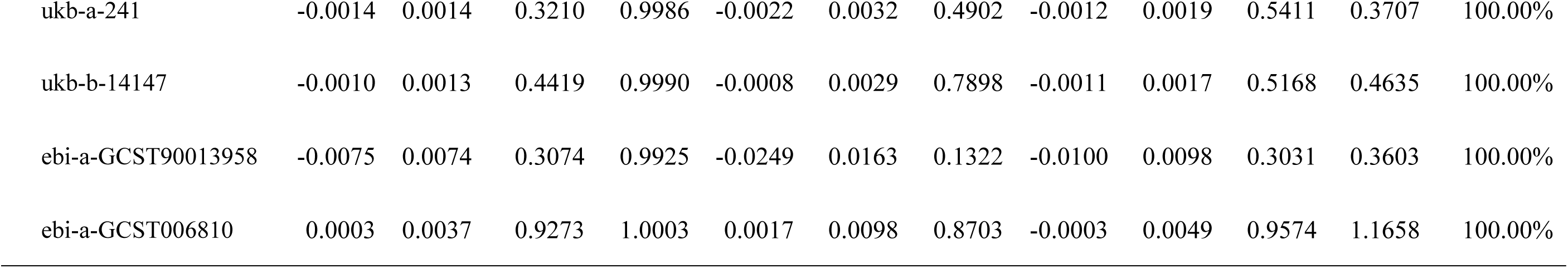
Summary of MR analyses with three regression approaches.

Our results showed a significant negative causal association between anti-*T. gondii* IgG levels and bipolar, that is, a higher anti-*T. gondii* IgG level reduces the risk of bipolar (Table 1). This contradicts the common hypothesis^15^. However, the significant results were driven by a single dataset ieu-b-5110. We did a colocalization analysis (data not shown) and the results support the hypothesis that only the exposure has a genetic association in the SNPs of the region with a posterior probability of 0.99. It suggests this causal association is likely a false positive. Besides the meta-analysis on anti-*T. gondii* IgG levels with bipolar disorder, the rest of meta-analyses showed there was no significant pooled causal effect of *T. gondii* infection on bipolar disorder and Schizophrenia (95% CIs of ORs all include 1), with a statistical power of ∼100% (S3-S4 Table, S38 Figure).

We also conducted reverse MR analysis to assess the causal impacts of psychiatric disorders or risk-taking behavior on *T. gondii* infection. In the context of homogeneous MR analyses, no significant effects were detected (S9-S12 Table).

### Assessing effects of *T. gondii* infection (anti-*T. gondii* IgG seropositivity) on psychiatric disorders

During two MR analyses with addiction as the outcome, the SNP rs4356554 was detected as an outlier in the GWAS dataset ukb-d-20552_2 through the MR-PRESSO outlier test (S5) and was subsequently deleted from the filtered IVs. The association of *T. gondii* infection with addiction did not achieve statistical significance in IVW regression model (Table 1). Neither the GWAS dataset ukb-d-20552_2 with the removal of the outlier nor the GWAS dataset ukb-d-20415 exhibited heterogeneity or pleiotropy (S5 Table). The nonsignificant result for the GWAS dataset ukb-d-20415 may be due to the very low power of 2.80% (Table 1), yet it is not the case for the GWAS dataset ukb-d-20552_2 as its statistical power in the two-sample MR analyses was ∼100% (Table 1).

In the four MR analyses with bipolar disorder as the outcome, MR-PRESSO outlier tests revealed the SNP rs147285933 as an outlier of IVs in the GWAS dataset ieu-b-41 (S5 Table), and it was deleted subsequently. IVW regression revealed no significant association between *T. gondii* infection and bipolar disorder (Table 1). Heterogeneity tests and MR-Egger intercept tests showed no evidence of heterogeneity or pleiotropy in the four MR analyses (S5 Table). Three MR analyses with the outcome GWAS datasets (ebi-a-GCST003724, ieu-a-801, and ieu-b-5110) had sufficient statistical power (>80%). However, the statistical power of MR analysis with the outcome GWAS dataset ieu-b-41 was low (31.70%) (Table 1).

With no outliers detected by MR-PRESSO, the association of *T. gondii* infection with obsessive-compulsive disorder was not statistically significant according to the IVW model (Table 1). There was no evidence of heterogeneity or pleiotropy in this two-sample MR analysis (S5 Table), and this analysis had satisfactory statistical power (>80%) (Table 1).

Among the seven MR analyses investigating the relation between *T. gondii* infection and Schizophrenia, the SNP rs7727080 in the GWAS dataset ieu-b-5102 was identified as an outlier via the MR-PRESSO outlier test. IVW regression indicated no statistically significant effects existed in the entire six MR analyses (Table 1). No heterogeneity and pleiotropy were found in two relevant tests (S5 Table). The statistical power of seven two-sample MR analyses was sufficient to detect effects (>80%) (Table 1).

Overall, in leave-one-out analyses with sensitivity tests, no SNP exhibited a dominant influence on the estimated effects across each MR analysis involving the outcomes of the four psychiatric disorders (S1-S14 Figure D. Leave-one-out Forest plot).

### Assessing the effects of *T. gondii* infection (anti-*T. gondii* IgG seropositivity) on risk-taking behavior

Four summarized GWAS datasets on risk-taking behavior were included as the outcome of our study. No outliers were identified in the four two-sample MR analyses based on MR-PRESSO tests. IVW regression yielded nonsignificant estimates for the effect of *T. gondii* infection on risk-taking behavior (Table 1). Heterogeneity and MR-Egger intercept tests confirmed the absence of heterogeneity and pleiotropy in all four two-sample MR analyses (S5 Table). The statistical power of all four MR analyses for the outcome of risk-taking behavior was consistently sufficient (>80%) (Table 1). Leave-one-out analyses with sensitivity tests revealed that the effect estimation was not influenced by any dominant SNP (S15-S18 Figure D. Leave-one-out Forest plot).

### Assessing effects of *T. gondii* infection (anti-*T. gondii* IgG levels) on psychiatric disorders

During two MR analyses with anti-*T. gondii* IgG levels as the exposure, no outlier SNPs were detected through the MR-PRESSO outlier test for the outcome addiction. The association of *T. gondii* infection with addiction did not achieve statistical significance in IVW regression model (Table 1). Neither the GWAS dataset ukb-d-20552_2 nor the GWAS dataset ukb-d-20415 exhibited heterogeneity or pleiotropy (S6 Table). The statistical power in these two two-sample MR analyses was greater than 80% (Table 1).

In four MR analyses with bipolar disorder as the outcome, MR-PRESSO outlier tests revealed the SNP rs60385209 as an outlier of IVs in the GWAS dataset ieu-b-41 (S6 Table), and it was deleted subsequently. IVW regression showed no significant positive association between anti-*T. gondii* IgG levels and bipolar disorder (Table 1). Please note that one of the dataset (ieu-b-5110) did suggest that there is a significant negative association between anti-*T. gondii* IgG levels and bipolar disorder. However, in contrast to the common hypothesis that higher anti-*T. gondii* IgG levels are associated with higher bipolar risk^36,37^, and the result suggests the opposite. Heterogeneity tests and MR-Egger intercept tests showed no evidence of heterogeneity or pleiotropy in the four MR analyses (S6 Table). All four two-sample MR analyses with the outcome of bipolar disorder had sufficient statistical power (>80%) (Table 1).

With no outliers detected by MR-PRESSO, the association of anti-*T. gondii* IgG levels on obsessive-compulsive disorder were not statistically significant according to the IVW model (Table 1). There was no evidence of heterogeneity or pleiotropy in this two-sample MR analysis (S6 Table), and this TWO-SAMPLE MR analysis had satisfactory statistical power (>80%) (Table 1).

Among the seven MR analyses investigating the relation between anti-*T. gondii* IgG levels on Schizophrenia, two SNPs, rs114230280 and rs1264457, in the GWAS dataset ieu-b-42 were identified as outliers via the MR-PRESSO outlier test. IVW regression indicated no statistically significant causal effects existed in the entire six MR analyses (Table 1). No heterogeneity and pleiotropy were found in two relevant tests (S6 Table). The statistical power of seven two-sample MR analyses was sufficient to detect effects (>80%) (Table 1).

Overall, in leave-one-out analyses with sensitivity tests, no SNP exhibited a dominant influence on the estimated effects across each MR analysis involving the outcomes of the four psychiatric disorders (S19-S32 Figure D. Leave-one-out Forest plot).

#### Assessing the effects of *T. gondii* infection (anti-*T. gondii* IgG levels) on risk-taking behavior

Four summarized GWAS datasets on risk-taking behavior were included as the outcome of our study. One outlier, rs77236966, was identified in the GWAS data ebi-a-GCST90013958 based on MR-PRESSO tests (S6 Table). IVW regression yielded nonsignificant estimates for the effect of anti-*T. gondii* IgG levels on risk-taking behavior (Table 1). Heterogeneity and MR-Egger intercept tests confirmed the absence of heterogeneity and pleiotropy in all four two-sample MR analyses (S6 Table). The statistical power of all four MR analyses for the outcome of risk-taking behavior was consistently sufficient (>80%) (Table 1). Leave-one-out analyses with sensitivity tests revealed that the effect estimation was not influenced by any dominant SNP (S33-S36 Figure D. Leave-one-out Forest plot).

#### Meta-analysis with multiple two-sample MR studies

To further increase the power to detect the potential causal impact of *T. gondii* infection (anti-*T. gondii* IgG seropositivity & levels), we conducted a meta-analysis for the outcome with multiple independent GWAS datasets in two-sample MR analyses, including bipolar disorder and Schizophrenia.

For the multiple two-sample MR studies with the exposure anti-*T. gondii* IgG seropositivity (GWAS data ebi-a-GCST90006925), a fixed-effects model demonstrated that there was no significant pooled causal effect of *T. gondii* infection on bipolar disorder and Schizophrenia, with a statistical power of ∼100% (S37 Figure A, B & C).

For the multiple two-sample MR studies with the exposure anti-*T. gondii* IgG levels (GWAS data ieu-b-4910), a fixed-effects model demonstrated that there was no significant positive pooled causal effect of *T. gondii* infection on bipolar disorder and Schizophrenia, with a statistical power of ∼100% (S37 Figure D, E & F).

Moreover, Cochran’s *Q* test revealed that none of the four meta-analyses in our study exhibited significant heterogeneity (S3-S4 Table). The Higgins *I*^2^ index statistics in meta-analyses for two-sample MR studies consistently approached 0%, indicating homogeneity among different two-sample MR studies (S3-S4 Table). The statistics τ^2^ for each meta-analysis were zero within a tiny standard error, signifying no variability in true effect sizes across different two-sample MR studies (S3-S4 Table). It was confirmed through leave-one-out analysis with sensitivity tests that the overall causal effect estimation was not influenced by any dominant two-sample MR analysis (S7-S8 Table). Importantly, each meta-analysis in our study demonstrated sufficient statistical power (∼100%) to detect the pooled causal effect for every exposure-outcome path.

### Reverse MR analyses

We also conducted reverse MR analysis to assess the causal impacts of psychiatric disorders or risk-taking behavior on *T. gondii* infection (anti-*T. gondii* IgG seropositivity & anti-*T. gondii* IgG levels). In the context of homogeneous MR analyses, no significant effects were detected in the reverse path of the MR (S9-S12 Table).

## Discussion

This study represents the first comprehensive analysis systematically investigating the causal effect of *T. gondii* infection (anti-*T. gondii* IgG seropositivity and levels) on four primary psychiatric disorders and risk-taking behavior. The findings indicate that exposure to *T. gondii* infection did not exhibit a statistically significant positive causal effect on the outcomes of psychiatric disorders (including addiction, bipolar disorder, obsessive-compulsive disorder, and Schizophrenia) or risk-taking behavior. This study provides crucial insights into the relationship between *T. gondii* infection and human psychopathology, emphasizing the need to re-evaluate existing hypotheses in this domain and the necessity of using robust analytical methods such as MR to decipher the intricate interplay between infectious diseases and mental health.

In our study, a relaxed significance threshold (*p* < 1×10^-5^ and *p* < 5×10^-5^) was applied to identify instrumental variants for several exposures. Additional statistical verification and testing of the selected SNPs were conducted to ensure the validity and robustness of the strong instrumental instruments in accordance with current MR analysis guidelines. To mitigate potential weak-IVs bias introduced by the relaxed threshold, we evaluated instrument strength for each selected SNP, all of which demonstrated adequate relevance (individual *F*-statistics > 16; mean *F*-statistics > 19) (S1-S2 Table). Both pleiotropy tests, including weighted median (Table 2.), MR-Egger test (S5-S6 Table), and MR-PRESSO test (S5-S6 Table), and sensitivity analyses, such as heterogeneity statistics (Cochran’s Q) (S5-S6 Table), and leave-one-out test (S2-S37 Figure D ), consistently supported the robustness of screened SNPs in our findings of MR analyses. Although applying a relaxed selection threshold may theoretically increase the risk of false positive instruments, the uniformly strong *F*-statistics, sufficient evidence of statistical tests (heterogeneity tests & sensitivity analysis), and biological coherence verification (pleiotropy tests) collectively reinforce the credibility of our conclusions.

**Table 2.**
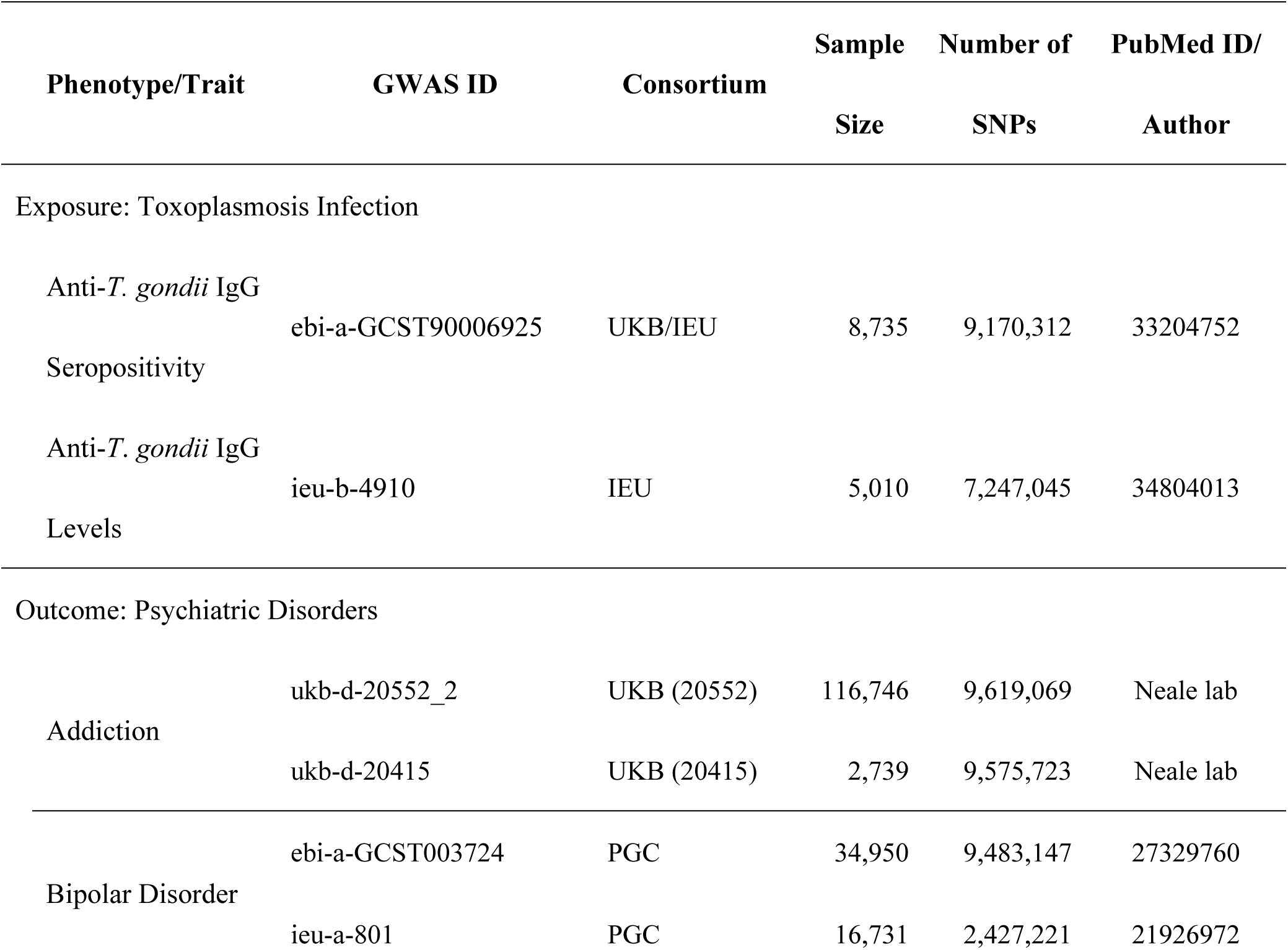
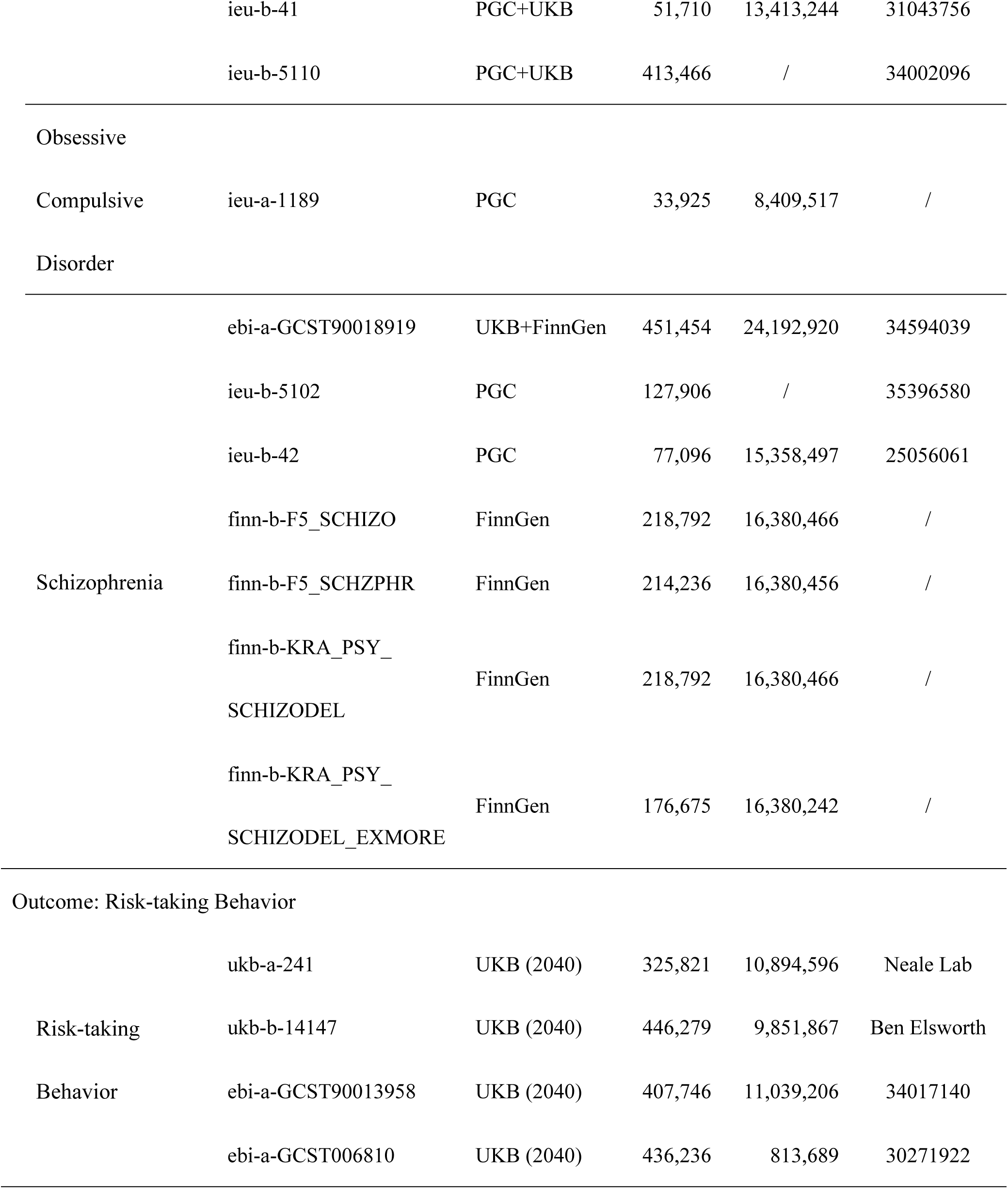
Summary of GWAS datasets on exposure and outcomes.

To aid in the interpretation of null findings, we examined the confidence intervals (CIs) of the causal estimates in our meta-analyses to determine the range of effect sizes that could be reasonably ruled out. For the analysis examining anti-T. gondii IgG seropositivity as the exposure and bipolar disorder as the outcome, the upper bound of the 95% CI for the odds ratio (OR = 0.951, 95% CI: [0.844, 1.073]; see S3 Table) suggests that a causal effect increasing the odds of bipolar disorder by more than 7.3% can be reasonably excluded. Similarly, for the association between anti-T. gondii IgG seropositivity and schizophrenia, the meta-analysis (OR = 1.020, 95% CI: [0.887, 1.185]; see S3 Table) rules out a causal effect that increases the odds of schizophrenia by more than 18.5%. Furthermore, for the analysis using anti-T. gondii IgG levels as the exposure and schizophrenia as the outcome, the results (OR = 1.010, 95% CI: [0.980, 1.030]; see S4 Table) indicate that a causal effect increasing the odds of schizophrenia by more than 3% can also be reasonably excluded. In other words, although we did not observe statistically significant associations, the results of meta-analysis provide sufficient evidence against the presence of moderate-to-large causal effects.

Our research is subject to several limitations. First, our MR analyses were conducted exclusively in European populations, especially in the UK and Finn populations. Caution must be exercised in generalizing our findings to other populations. Second, even though selecting SNPs as IVs adheres to stringent criteria grounded in the three core assumptions of MR analysis, we cannot definitively exclude the possibility that some genetic variants may manifest unknown pleiotropic effects. This potential scenario could pose a challenge to the fulfillment of the third assumption in MR analysis. Finally, the exposure measurement in this study may not cover the entire spectrum of the impact of *T. gondii* infection. For example, they cannot distinguish the type of *T. gondii* infection: congenital, acute, chronic, or latent. They also do not directly measure the duration of infection. Therefore, we cannot exclude the possibility that prolonged chronic infection may have a causal impact on psychopathology.

## Materials & Methods

### Study design

According to Mendel’s law of independent assortment, parental alleles (gene variants) for different traits segregate, or assort, into gametes independently of one another during gamete formation. This process of Mendelian randomization is analogous to the randomization employed in randomized controlled trials^9^. This study leveraged SNPs in GWAS datasets as instrumental variables (IVs) to explore the potential causal association between the exposure of interest and the outcome. The entire study design for the two-sample MR analysis was developed, as illustrated in S1 Figure. Toxoplasmosis infection served as the exposure variable, while the occurrence of four psychiatric disorders and one risk-taking behavior was considered outcome variables.

### Data resource

Genome-wide association study summary-level statistics for two different measures of Toxoplasmosis infection (anti-*T. gondii* IgG seropositivity, and anti-*T. gondii* IgG levels), four types of psychiatric disorders (addiction, bipolar disorders, obsessive-compulsive disease, and Schizophrenia) and risk-taking behavior were obtained from the UK Biobank (UKB) (https://biobank.ctsu.ox.ac.uk/), the Integrative Epidemiology Unit OpenGWAS project (IEU) (https://gwas.mrcieu.ac.uk/), the Finnish Genomics project (FinnGen) (https://public-metaresults-fg-ukbb.finngen.fi/) and the Psychiatric Genomics Consortium (PGC) (https://www.med.unc.edu/pgc/). To avoid the bias of population heterogeneity, all participants in the summarized GWAS data were solely from European populations. A total of 18 summary-level GWAS datasets were utilized in our MR analyses (Table 2).

### Instrumental variable selection

The selection of IVs is a crucial step before conducting two-sample MR analyses. This is because SNPs, as exposure proxies, must ensure valid causal inference. Therefore, the SNPs included in this research should satisfy three core assumptions for IVs: i) The relevance assumption: the single nucleotide polymorphisms (SNPs) used as instrumental variables (IV) should be strongly correlated with the exposure of interest; ii) The exclusion-restriction assumption: the IVs should exclusively influence the outcome through the exposure and not exhibit pleiotropic effects; and iii) The independence assumption: the IVs should not be associated with any confounding factors related to the exposure-outcome relationship. First, the SNPs strongly associated with anti-*T. gondii* IgG seropositivity and anti-*T. gondii* IgG levels were extracted at a genome-wide significance threshold of *p* < 1×10^-5^ and *p* < 5×10^-5^, respectively, to ensure that the selected IVs fulfilled the assumptions. The relaxation of the genome-wide significance threshold (5×10^-8^) was implemented to ensure an adequate sample size of SNPs for solid two-sample MR analyses^16^. Second, applying linkage disequilibrium (LD) clumping with PLINK (v1.9)^17^ ensured the selection of independent SNPs (LD threshold: LD *r*^2^ < 0.001, LD region width > 10,000 kb), in which sequencing data from the 1000 Genomes Project Phase 3 were used to estimate LD. Then, the *F*-statistics, defined as ((*N*-*k*-1)/*k*) · (R^2^ /((1-R^2^)) (*N*: sample size of participants; *k*: number of independent SNPs; R^2^: variance in exposure explained by SNPs)^18^, was calculated to assess the strength of each filtered SNP. SNPs with *F*-statistics less than 10 were considered weak IVs and were excluded.

To eliminate the impact of horizontal pleiotropy bias and ensure adherence to the second assumption, we rigorously validated all screened independent SNPs by using the PhenoScanner online database of genotype-phenotype associations (http://www.phenoscanner.medschl.cam.ac.uk) (date checked Dec. 2023). This verification process involved assessing whether the selected SNPs were associated with secondary phenotypes, considering a stringent significance threshold (*p-value* < 5×10^-8^). Ultimately, a set of 25 and 76 genome-wide significant SNPs serving as IVs strongly associated with the exposure to *T. gondii* infection was selected carefully through screening of the summary-level GWAS dataset ebi-a-GCST90006925 and ieu-b-4910 (see Supplementary Table 4 & 10). Before conducting two-sample Mendelian randomization, palindromic SNPs in two sets of IVs with minor allele frequencies (MAFs) exceeding 0.42 or SNPs with allele mismatches were excluded during the data harmonization process.

### Statistical analysis

Two-sample MR analyses, investigating causal associations between an exposure (risk factor) and an outcome using genetic data from two independent samples or datasets, were subsequently conducted with two R packages, "TwoSampleMR"^19^ and "MR-PRESSO"^20^. The priori statistical power for MR analyses was calculated via the Shiny web application (https://sb452.shinyapps.io/power/)^21^.

To evaluate the causal effect of *T. gondii* infection (anti-*T. gondii* IgG seropositivity and anti-*T. gondii* IgG levels) on four different psychiatric disorders and risk-taking behavior, two-sample MR analyses were implemented by using the inverse variance weighting (IVW) approach^22^, the MR-Egger regression approach^23^, the MR-Pleiotropy RESidual Sum and Outlier (MR-PRESSO) approach^20^ and the weighted median approach^24^. The various IV regression methods used in two-sample MR analysis involve distinct model assumptions, and our study leveraged different approaches, each of which plays a unique role based on these assumptions.

The IVW regression method provided robust and accurate estimates of causal effects under the assumption of no horizontal pleiotropy. However, the MR-Egger method and the weighted median method were used to conduct MR analysis, assuming the presence of pleiotropy in >50% of the SNPs^23^ (Bowden et al., 2015) and <50% of the SNPs^24^, respectively. The MR-PRESSO method not only provides a corrected estimate of causal effects on pleiotropy but also allows statistical tests to recognize and delete outlier and pleiotropic SNPs in MR analyses under the assumption of pleiotropy^20^. Consequently, the primary MR analyses relied on the IVW method, provided that no horizontal pleiotropy was detected in the sensitivity analyses. In the presence of horizontal pleiotropy, the MR-Egger regression method can provide more valid Mendelian randomization estimates than the IVW method^25^. When heterogeneity is identified among the IVs without the presence of pleiotropy, employing the weighted median method can provide robust estimates in two-sample MR analysis^24^.

To assess the reliability of the results of a two-sample MR analysis, it is crucial to conduct a comprehensive sensitivity analysis, which helps to check and address underlying horizontal pleiotropy and heterogeneity. The MR-Egger intercept test can detect potential violations of the second assumption regarding horizontal pleiotropy with a *p-value* less than 0.05. Furthermore, the MR-PRESSO method can reveal the existence of overall horizontal pleiotropic effects by implementing global tests and checking for horizontal pleiotropic outliers by performing outlier tests. After excluding the outlier SNP with an individual *p-value* less than 0.05 in the outlier test, the MR-PRESSO global and outlier tests were repeated for the remaining SNPs until the global test yielded statistical nonsignificance (p value>0.05). The subsequent two-sample MR analysis was carried out on the remaining SNPs. Cochran’s *Q* test detected heterogeneity among SNPs in the IVW regression, where a Q *p-value* above 0.05 indicated that the null hypothesis of no heterogeneity could not be rejected^26^. Leave-one-out analyses with sensitivity tests were employed as a useful tool for detecting genetic variants that dominate the causal effect estimate. The removal of such SNPs can enhance the robustness of the findings in two-sample MR analyses. The reverse two-sample MR analyses were performed following the same procedure as those described above.

For meta-analysis, we first checked the overlap in the samples included in these studies for outcomes involving multiple GWAS datasets. Specifically, two relevant GWAS datasets related to psychiatric disorder addiction are focused on behavioral and miscellaneous addictions (ukb-d-20552_2) and ongoing addiction to alcohol (ukb-d-20415). Therefore, combining the two studies in meta-analysis can be problematic. In four GWAS studies on bipolar disorder, ieu-b-41 (32 cohorts)^27^ was identified as a subset of ieu-b-5110 (57 cohorts)^28^, which also encompassed the cohort with bipolar disorder in the UK Biobank. When comparing the cohorts included in the remaining GWAS studies, the dataset ebi-a-GCST003724 (Northern Swedish GWAS)^29^, ieu-a-801 (Psychiatric GWAS Consortium Bipolar Disorder Working Group, 2011) and ieu-b-5110 were found to be mutually independent. In seven GWAS datasets on Schizophrenia, three datasets from the FinnGen database (finn-b-F5_SCHIZO, finn-b-KRA_PSY_SCHIZODEL, finn-b-KRA_PSY_SCHIZODEL_EXMORE) were focused on Schizophrenia or delusions, symptoms often associated with Schizophrenia. The dataset finn-b-F5_SCHZPHR was the only one purely related to Schizophrenia from the FinnGen database. The dataset ebi-a-GCST90018919^30^ harmonized cohorts from the UK Biobank and FinnGen database in the GWAS study of Schizophrenia. The cohorts covered in the dataset ieu-b-42 (Schizophrenia Working Group of the Psychiatric Genomics Consortium, 2014) were entirely included in the GWAS study with the dataset ieu-b-5102^31^. All GWAS datasets on risk-taking behavior were derived from cohorts in the UK Biobank database (field ID:2040) with overlap. Therefore, to ensure the independence of MR studies within the common outcome, three groups of independent GWAS datasets on bipolar disorder (ebi-a-GCST003724, ieu-a-801, ieu-b-5110) and Schizophrenia (subset (i): ebi-a-GCST90018919, ieu-b-5102; subset (ii): ieu-b-5102, finn-b-F5_SCHZPHR) were utilized for conducting meta-analysis to obtain pooled causal effects. The pooled estimate of causality was derived through meta-analysis using the R package "metafor"^32^. Cochran’s *Q* test and Higgins *I^2^* index statistics were employed to assess heterogeneity among multiple two-sample MR analyses for each outcome. If the *p-value* in the heterogeneity test exceeded 0.05 and the *I*^2^ index was less than 50%, suggesting homogeneity among two-sample MR analyses, the fixed-effects model was deemed appropriate. The fixed-effects model assumes that effect sizes are consistent across studies and estimates a weighted average of their true values^33^. Conversely, the random effects model was considered more suitable if heterogeneity was detected among two-sample MR analyses (*p-value* < 0.05 or *I*^2^ index statistics ≥ 50%). The random effects model assumes that the study effects are variable and that the studies represent a random sample from a larger population of similar studies^33–35^. The statistics tau-square (τ^2^), an estimate of the between-study variance, was also reported in our meta-analysis, representing the degree of variability in true effect sizes across studies beyond what would be expected owing to random sampling error alone. The greater the value of τ^2,^ the more between-study heterogeneity in the true effect sizes across different two-sample MR studies. Furthermore, Egger’s test was used to evaluate publication bias. A sensitivity test and leave-one-out analysis were used in the meta-analysis to assess the overall results’ robustness and investigate the influence of individual two-sample MR analysis on the summarized causal estimate. These analyses helped identify whether specific studies heavily influence the findings and whether the results are consistent across different GWAS datasets within a given exposure-outcome pathway with causality.

The post-hoc statistical power for MR analyses was calculated via the Shiny web application (https://sb452.shinyapps.io/power/), based on the effect sizes of the causal effect estimates obtained from the IVW regression method. Power calculations were performed using the Shiny web application (https://jason-griffin.shinyapps.io/shiny_metapower/) to gauge the statistical power of the meta-analysis.

## Author Contributions

Concept and design: Liu.

Acquisition, analysis, or interpretation of data: Pang, Liu.

Drafting of the manuscript: Pang, Liu.

Critical review of the manuscript for important intellectual content: All authors.

Statistical analysis: Pang.

Obtained funding: Liu.

Administrative, technical, or material support: Liu.

Supervision: Liu.

## Data availability statement

The data supporting the findings of this study are openly available in the Integrative Epidemiology Unit OpenGWAS project (IEU) at https://gwas.mrcieu.ac.uk/. Reference numbers are provided in Table 2 of the manuscript.

